# Awareness, support, and motivation surrounding the Canadian Dental Care Plan

**DOI:** 10.64898/2026.11.09.26367626

**Authors:** Giselle CO Pessoa, Thiago HL Baltus, Carol Youssef, Eleanor Reimer, Anil Menon, Nikhil Lockman, Robert J Schroth

## Abstract

**Objective:** The Canadian Dental Care Plan (CDCP) was introduced to provide dental insurance for low-income residents of Canada. The purpose of this study was to investigate CDCP awareness, support, alongside motivators to apply for the plan among a national sample of Canadian residents.

**Methods:** Data from a national cross-sectional public opinion survey conducted before CDCP implementation, on behalf of Health Canada’s Oral Health Branch, were analyzed. The survey included 2,250 participants from urban and rural/remote communities across Canada and assessed baseline awareness of the CDCP and barriers to dental care. Data were obtained from the Government of Canada Publications online archive. Descriptive and bivariate analyses were applied to evaluate associations among CDCP awareness, support, and motivators, with sociodemographic covariates. Statistical significance was set at *p* ≤ 0.05.

**Results:** Overall, 42.7% of respondents had heard of the CDCP, with lower awareness among females, younger adults, those with lower education, those employed, and those never married. Female parents and younger adults were more likely to have searched for CDCP information, most commonly through news and media (53.0%). Overall, 81.5% supported the CDCP, with greater support among those without dental insurance, with university education, and those from Atlantic Canada, British Columbia, Yukon, Northwest Territories, and Nunavut. Meanwhile, opposition was higher in the Prairie Provinces. Leading motivators for applying included the extent of coverage offered, prospect of a better dental health, and whether the enrollment process would be easy. Uninsured participants were significantly more likely to apply for the CDCP if they reported having a good understanding of the plan, lived close to an oral health professional, and had urgent dental needs.

**Conclusion:** Despite low pre-implementation awareness, pre-implementation support for the CDCP was high. Results can serve as a national baseline of CDCP awareness and support to measure changes in the public’s views of the CDCP, and its potential role in improving access to essential oral health care in Canada. Based on these observations, follow-up studies are essential to evaluate the program’s reach, development and uptake following implementation.

## 1. Introduction

Until recently, oral health care has not been part of public health care in Canada. Although the Canada Health Act was designed to “protect, promote and restore the physical and mental well-being of residents of Canada and to facilitate reasonable access to health services”, cost and socioeconomic circumstances used to restrict many Canadians from accessing dental care.^1^ Past surveys have identified that nearly one quarter of Canadians have avoided seeking dental care due to affordability barriers. ^2^ Unfortunately, the World Health Organization (WHO) has reported that dental expenses often result in families facing economic hardship^3^ and the FDI World Dental Federation agrees that treatment costs are too high, especially for marginalized and poor people.^4^

Despite lasting inconsistencies among equal access to dental care, financial resources, and disparities across provinces^5^, the Canadian government made a significant commitment to make oral health care more affordable for low-income families in 2022. As part of this pledge the Federal government introduced the Interim Canada Dental Benefit (CDB) for children <12 years of age from families with an adjusted household net income < $90,000 and no access to private dental insurance.^6–8^ The Interim CDB, which ran between October 2022 to June 2024, was a precursor to the Canadian Dental Care Plan (CDCP) program. The CDCP was launched in the spring of 2024 for low-income seniors but began accepting applications for children <18 years and adults with disabilities (based on valid disability tax credits) soon after. In 2025, the CDCP was expanded to include all adults aged 18 to 64 years,^9^ covering the needs of Canadian residents based on their Annual Family Net Income (AFNI): recipients with an AFNI lower than $70,000 have 100% of their dental expenses covered, those with an AFNI between $70,000 and 79,999 receive 60% coverage, while those with an AFNI between 80,000 and 89,999 receive 40% coverage.^9^

Given the importance of establishing the CDCP as a national oral health insurance program for lower-income residents of Canada, the purpose of this study was to investigate the awareness of and support for the CDCP along with motivators to apply for the plan among a national sample of residents of Canada.

## 2. Methods

This cross-sectional study analyzed data obtained from a 2023 online public opinion survey conducted from July 26, 2023 to August 23, 2023 by the Strategic Counsel on behalf of Health Canada’s Oral Health Branch.^10^ The survey was available online, while eligible participants from remote areas were approached by phone. Quota sampling was employed for the base sample (n= 2,150) to ensure broad representation of Canadian adults who met the eligibility requirements of the CDCP, and eligible families from remote communities were recruited by telephone (n=100). A total of 2,250 individuals, over 18 years of age, were included in the study, comprising the base sample. Due to oversampling from Atlantic Canada (includes New Brunswick, Newfoundland and Labrador, Nova Scotia, and Prince Edward Island) and the Prairies (includes Manitoba and Saskatchewan), a weighting scheme was used to normalize the distribution of the population by regions in Canada. The required sample size was calculated to achieve a +5% margin of error at a 95% confidence level for key variables, relative to the target population. More details of the methods used can be found in the Canada Dental Benefit - Expanded Baseline Survey Final Report.^10^ Data were accessed from the Government of Canada Publications website, at: https://epe.bac-lac.gc.ca/100/200/301/pwgsc-tpsgc/por-ef/health/2024/014-23-e/014-23_Report.pdf.^11^

Institutional ethics approval from the University of Manitoba was deemed unnecessary, as this study used aggregated and anonymized data without personal identifiers. Individual characteristics (e.g., gender, age, access to insurance, etc.) were evaluated on a national level, with more specified evaluation using regional data. For variables with less than 5 respondents, values were suppressed or consolidated into other categories for confidentiality purposes.

Analyses were conducted for the overall sample (n=2,250), with additional analyses of data relating to parents of children under 12 years of age (n=298) and those parents of children 12 to 17 years (n=275). Outcome variables of interest included whether respondents had heard about the CDCP, whether they had searched for information about the CDCP in the last 3 months, their motivators to apply for the CDCP, as well as their support for the program. Sociodemographic covariates of interest included participant gender, age, education level, employment status, marital status, area of residence (urban, rural or remote), region of Canada, their dental insurance situation, and whether they had children under 12 years, 12 to 17 years, 18 or older, or had no children.

Statistical analyses included descriptive statistics (e.g., frequencies, means, and standard deviations (SD)) and bivariate analyses (Z-test, 95% confidence interval). A p-value <0.05 was considered significant.

## 3. Results

A total of 2,250 individuals participated with the majority being female (51.7%), under 45 years of age, married or in common-law relationships (44.5%), and lacking dental insurance (69.8%) (Table 1). Most participants had completed post-secondary education (67.1%) and lived in urban centres (82.6%).

**Table 1.**
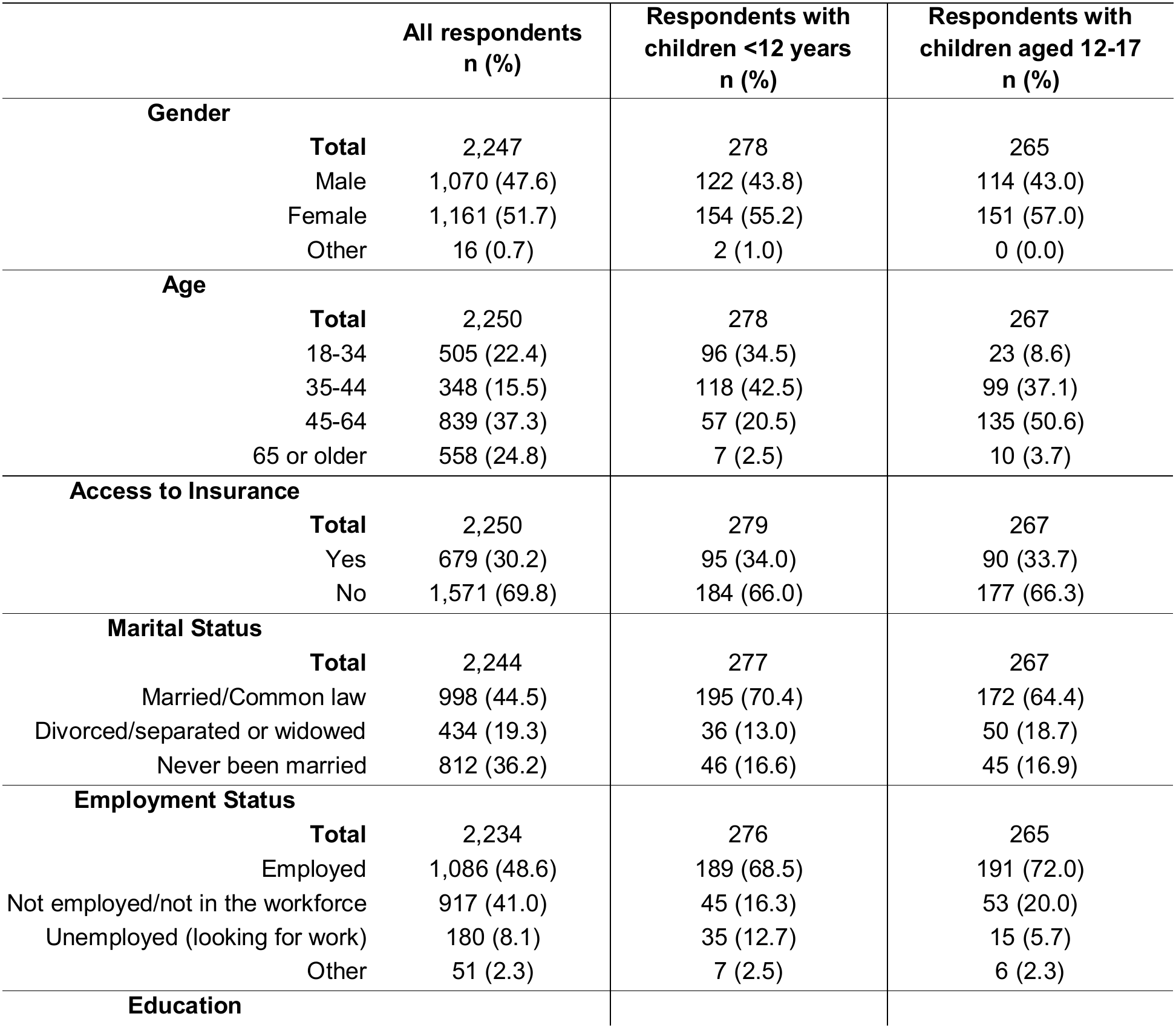
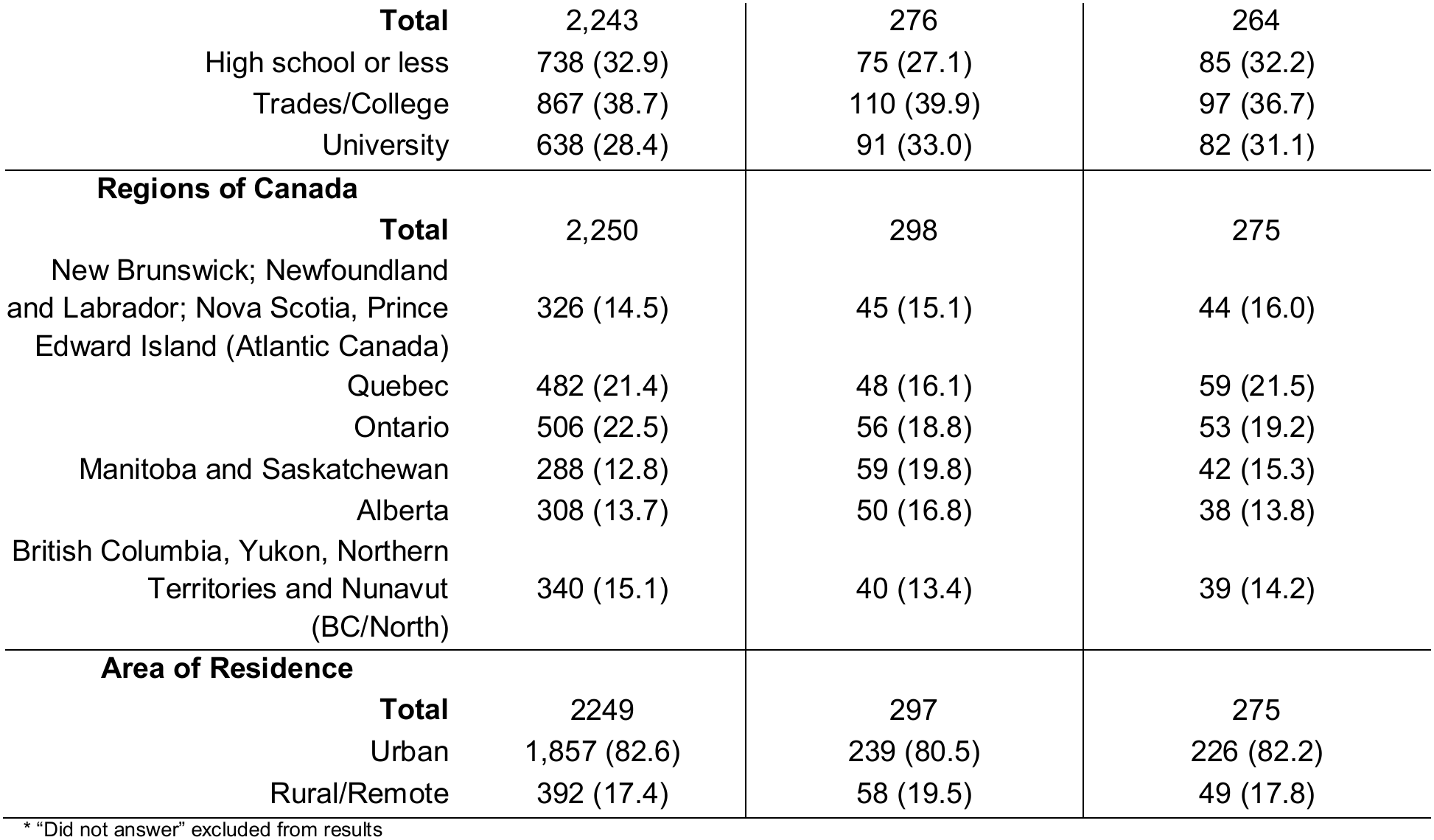
Characteristics of Participants.

|  | All respondents<br>n (%) | Respondents with children <12 years<br>n (%) | Respondents with children aged 12-17<br>n (%) |
|---|---|---|---|
| **Gender** |  |  |  |
| **Total** | 2,247 | 278 | 265 |
| Male | 1,070 (47.6) | 122 (43.8) | 114 (43.0) |
| Female | 1,161 (51.7) | 154 (55.2) | 151 (57.0) |
| Other | 16 (0.7) | 2 (1.0) | 0 (0.0) |
| **Age** |  |  |  |
| **Total** | 2,250 | 278 | 267 |
| 18-34 | 505 (22.4) | 96 (34.5) | 23 (8.6) |
| 35-44 | 348 (15.5) | 118 (42.5) | 99 (37.1) |
| 45-64 | 839 (37.3) | 57 (20.5) | 135 (50.6) |
| 65 or older | 558 (24.8) | 7 (2.5) | 10 (3.7) |
| **Access to Insurance** |  |  |  |
| **Total** | 2,250 | 279 | 267 |
| Yes | 679 (30.2) | 95 (34.0) | 90 (33.7) |
| No | 1,571 (69.8) | 184 (66.0) | 177 (66.3) |
| **Marital Status** |  |  |  |
| **Total** | 2,244 | 277 | 267 |
| Married/Common law | 998 (44.5) | 195 (70.4) | 172 (64.4) |
| Divorced/separated or widowed | 434 (19.3) | 36 (13.0) | 50 (18.7) |
| Never been married | 812 (36.2) | 46 (16.6) | 45 (16.9) |
| **Employment Status** |  |  |  |
| **Total** | 2,234 | 276 | 265 |
| Employed | 1,086 (48.6) | 189 (68.5) | 191 (72.0) |
| Not employed/not in the workforce | 917 (41.0) | 45 (16.3) | 53 (20.0) |
| Unemployed (looking for work) | 180 (8.1) | 35 (12.7) | 15 (5.7) |
| Other | 51 (2.3) | 7 (2.5) | 6 (2.3) |
| **Education** |  |  |  |
| **Total** | 2,243 | 276 | 264 |
| High school or less | 738 (32.9) | 75 (27.1) | 85 (32.2) |
| Trades/College | 867 (38.7) | 110 (39.9) | 97 (36.7) |
| University | 638 (28.4) | 91 (33.0) | 82 (31.1) |
| **Regions of Canada** |  |  |  |
| **Total** | 2,250 | 298 | 275 |
| New Brunswick; Newfoundland and Labrador; Nova Scotia, Prince Edward Island (Atlantic Canada) | 326 (14.5) | 45 (15.1) | 44 (16.0) |
| Quebec | 482 (21.4) | 48 (16.1) | 59 (21.5) |
| Ontario | 506 (22.5) | 56 (18.8) | 53 (19.2) |
| Manitoba and Saskatchewan | 288 (12.8) | 59 (19.8) | 42 (15.3) |
| Alberta | 308 (13.7) | 50 (16.8) | 38 (13.8) |
| British Columbia, Yukon, Northern Territories and Nunavut (BC/North) | 340 (15.1) | 40 (13.4) | 39 (14.2) |
| **Area of Residence** |  |  |  |
| **Total** | 2249 | 297 | 275 |
| Urban | 1,857 (82.6) | 239 (80.5) | 226 (82.2) |
| Rural/Remote | 392 (17.4) | 58 (19.5) | 49 (17.8) |

\* “Did not answer” excluded from results

Among respondents with children <12 years of age, 55.2% were female, between 35-44 years of age (42.4%), married or in common law relationships (70.4%), lacked access to dental insurance (66.0%) and lived in urban centres (80.5%). Meanwhile, among respondents with children aged 12-17, 57.0% were female, between 45-64 years of age (50.6%), married or in common-law relationships (64.4%), lacked access to dental insurance (66.3%), and lived in an urban area (82.2%).

Overall, 42.7% of respondents had heard of the CDCP. Several covariates were significantly associated with having heard about the program (Table 2). Males were significantly more likely to have heard of the CDCP than females (45.6% vs. 40.4%), while those aged 65+ years were also significantly more likely to have heard about the plan than all other age groups. Meanwhile, participants aged 35-44 and 45-64 were significantly more likely to have heard about the CDCP than those 18-34 years of age. Respondents with an education level ≤ high school were significantly less likely to have heard of the CDCP than those who completed college/trades and university (34.2% vs. 44.9% and 49.5%, respectively). Those not employed/not in the workforce were significantly more aware of the CDCP than employed and unemployed (looking for work) participants (48.3% vs. 39.7% and 32.2%, respectively). Divorced/separated or widowed respondents were more likely to have heard of the CDCP (49.9%) than those never married as well as those married or in common-law relationships (49.9% vs. 38.1% and 43.4%, respectively).

**Table 2.** Awareness of the new Government Program (Canadian Dental Care Plan)

|  | Heard about the new Government Program n (%) |  |  |  | Searched for information about the new dental program in the last 3 months n (%) |  |  |
|---|---|---|---|---|---|---|---|
|  | Total | Yes | No | Don’t know | Total | Yes | No |
| **Total overall** | **2250** | **961 (42.7)** | **1189 (52.9)** | **99 (4.4)** | **977** | **307 (31.9)** | **654 (68.1)** |
| **Gender** |  |  |  |  |  |  |  |
| Male (A) | 1,070 | **488 (45.6)<sup>B</sup>** | 538 (50.3) | 44 (4.1) | 487 | 139 (28.5) | **348 (71.5)<sup>B</sup>** |
| Female (B) | 1,161 | 469 (40.4) | **638 (55.0)<sup>A</sup>** | 54 (4.6) | 469 | **165 (35.2)<sup>A</sup>** | 304 (64.8) |
| Other (C) | 16 | 5 (32.3) | 10 (62.5) | 1 (6.2) | 5 | 3 (60.0) | 2 (40.0) |
| **Access to Dental Insurance** |  |  |  |  |  |  |  |
| Yes (D) | 679 | 286 (42.1) | 364 (53.6) | 29 (4.3) | 286 | 78 (27.3) | 208 (72.7) |
| No (E) | 1,570 | 675 (43.0) | 825 (52.5) | 70 (4.5) | 675 | 229 (33.9) | 446 (66.1) |
| **Age** |  |  |  |  |  |  |  |
| 18-34 (F) | 505 | 155 (30.7) | **335 (66.3)<sup>G,H,I</sup>** | 15 (3.0) | 155 | **70 (45.2)<sup>H,I</sup>** | 85 (54.8) |
| 35-44 (G) | 349 | **138 (39.5)<sup>F</sup>** | **186 (53.3)<sup>I</sup>** | **25 (7.2)<sup>F,H</sup>** | 137 | **57 (41.6)<sup>H,I</sup>** | 80 (58.4) |
| 45-64 (H) | 838 | **379 (45.2)<sup>F</sup>** | **426 (50.9)<sup>I</sup>** | 33 (3.9) | 379 | **115 (30.3)<sup>I</sup>** | **264 (69.7)<sup>F,G</sup>** |
| 65+ (I) | 558 | **289 (57.8)<sup>F,G,H</sup>** | 242 (43.4) | 27 (4.8) | 289 | 64 (22.1) | **225 (77.9)<sup>F,G,H</sup>** |
| **Education** |  |  |  |  |  |  |  |
| High school or less (J) | 738 | 252 (34.2) | **449 (60.8)<sup>K,L</sup>** | 37 (5.0) | 252 | 67 (26.6) | 185 (73.4) |
| College/Trades (K) | 867 | **389 (44.9)<sup>J</sup>** | **447 (51.5)<sup>L</sup>** | 31 (3.6) | 389 | 131 (33.7) | 258 (66.3) |
| University (L) | 638 | **316 (49.5)<sup>J</sup>** | 291 (45.6) | 31 (4.9) | 316 | 106 (33.5) | 210 (66.5) |
| **Employment status** |  |  |  |  |  |  |  |
| Employed (M) | 1,085 | 431 (39.7) | **608 (56.0)<sup>O</sup>** | 46 (4.3) | 432 | **162 (37.5)<sup>O</sup>** | 270 (62.5) |
| Unemployed (looking for work) (N) | 180 | 58 (32.2) | **112 (62.2)<sup>O</sup>** | 10 (55.6) | 58 | 22 (37.9) | 36 (62.1) |
| Not employed/not in workforce (O) | 917 | **443 (48.3)<sup>M,N</sup>** | 433 (47.2) | 41 (4.5) | 443 | 110 (24.8) | **333 (75.2)<sup>M,P</sup>** |
| Other (P) | 51 | 21 (41.1) | 29 (56.9) | 1 (2.0) | 22 | **11 (50.0)<sup>O</sup>** | 11 (50.0) |
| **Marital status** |  |  |  |  |  |  |  |
| Never Married (Q) | 811 | 309 (38.1) | **466 (57.5)<sup>R,S</sup>** | 36 (4.4) | 310 | 85 (27.4) | 225 (72.6) |
| Married or Common-Law (R) | 999 | **434 (43.4)<sup>Q</sup>** | 520 (52.1) | 45 (4.5) | 434 | 148 (34.1) | 286 (65.9) |
| Divorced/separated or Widowed (S) | 435 | **217 (49.9)<sup>Q,R</sup>** | 200 (46.0) | 18 (4.1) | 216 | 74 (34.3) | 142 (65.7) |
| **Parent of children** |  |  |  |  |  |  |  |
| <12 years (T) | 278 | 130 (46.8) | 138 (49.6) | 10 (3.6) | 130 | **73 (56.2)<sup>V,W</sup>** | 57 (43.8) |
| Age 12-17 (U) | 266 | **140 (52.6)<sup>V,W</sup>** | 118 (44.4) | 8 (3.0) | 141 | **67 (47.5)<sup>V,W</sup>** | 74 (52.5) |
| Age ≥18 (V) | 386 | 164 (42.5) | 201 (52.1) | 21 (5.4) | 164 | 55 (33.5) | **109 (66.5)<sup>T,U</sup>** |
| No children (W) | 1,435 | 595 (41.5) | **780 (54.3)<sup>U</sup>** | 60 (4.2) | 594 | 150 (25.3) | **444 (74.7)<sup>T,U</sup>** |
| **Regions of Canada** |  |  |  |  |  |  |  |
| New Brunswick; Newfoundland and Labrador; Nova Scotia, Prince Edward Island (Atlantic Canada) (X) | 326 | **151 (46.3)<sup>AB</sup>** | 165 (50.6) | 10 (3.1) | 151 | 53 (35.1) | 98 (64.9) |
| Quebec (Y) | 482 | 209 (43.4) | 254 (52.7) | 19 (3.9) | 209 | 63 (30.1) | 146 (69.9) |
| Ontario (Z) | 506 | 212 (41.9) | 269 (53.2) | 25 (4.9) | 212 | 69 (32.5) | 143 (67.5) |
| Manitoba and Saskatchewan (AA) | 288 | **138 (47.9)<sup>AB</sup>** | 138 (47.9) | 12 (4.2) | 138 | 46 (33.3) | 92 (66.7) |
| Alberta (AB) | 308 | 117 (38.0) | **175 (56.8)<sup>AA</sup>** | 16 (5.2) | 117 | 42 (35.9) | 75 (64.1) |
| British Columbia, Yukon, Northern Territories and Nunavut (BC/North) (AC) | 340 | 150 (44.1) | 176 (51.8) | 14 (4.1) | 150 | 43 (28.7) | 107 (71.3) |
| **Area of residence** |  |  |  |  |  |  |  |
| Urban (AD) | 1,857 | 808 (43.5) | 962 (51.8) | 87 (4.7) | 808 | 263 (32.5) | 545 (67.5) |
| Rural/Remote (AE) | 392 | 168 (42.8) | 215 (54.9) | 9 (2.3) | 168 | 52 (31.0) | 116 (69.0) |

Letters in uppercase indicate comparisons within the same column with a *p*-value <0.05
Unanswered questions were not included

Respondents from Manitoba and Saskatchewan and from Atlantic Canada were significantly more likely to have heard about the CDCP than those from Alberta. No associations were found between having heard of the program and whether respondents had access to dental insurance or whether they lived in urban or rural areas.

Associations between searching for the information about the program in the last 3 months and covariates are also reported in Table 2. Females were significantly more likely to have searched for information on the CDCP than males (35.2% vs. 28.5%). Those aged 18-34 and 35-44 were significantly more likely to have searched for information on the program than those aged 45-64 and those aged ≥ 65 years. Meanwhile, no associations were identified between searching for information on the CDCP in the past 3 months and respondents’ access to dental insurance, level of education, marital status, age of their children, area of residence, and region of Canada.

Information about where respondents heard about the CDCP are reported in Table 3. The most cited source was news and media outlets (53.5%), followed by social media (13.3%), advertising (12.1%), and family and friends (7.7%). Only 2.9% of respondents indicated having heard about the CDCP from oral health professionals. Respondents ≥ 65 years of age were significantly more likely to have heard of the CDCP from the news/media than all other age groups, and those not employed/not in the workforce were also more likely to have heard of the CDCP from the news media than those employed and those unemployed. Interestingly, parents of children ≥ 18 years of age were statistically more likely to have heard of the CDCP from the news or media than those with children < 12 years of age and those with children 12-17 years of age. Younger respondents (18-34) were significantly more likely to have learned significantly more about the program through social media when compared to those aged 45-64 and ≥ 65 years of age, while employed respondents were also statistically more likely to have learned about it on social media than those not employed/not in the workforce.

**Table 3.** Sources where respondents heard about the CDCP from, distributed by sociodemographic variables.

|  | **Total** | On the news/ media | From a healthcare provider | From an oral health professional | Friends or family | In a blog | On social media | Saw or heard an ad | CRA* website | Government (website, letter, email) |
|---|---|---|---|---|---|---|---|---|---|---|
| **Total overall** | **1,294** | **692 (53.5)** | **39 (3.0)** | **37 (2.9)** | **100 (7.7)** | **30 (2.3)** | **172 (13.3)** | **156 (12.1)** | **37 (2.9)** | **31 (2.3)** |
| **Gender** |  |  |  |  |  |  |  |  |  |  |
| Male (A) | 710 | **372 (52.4)<sup>B</sup>** | **26 (3.7)<sup>B</sup>** | **28 (3.9)<sup>B</sup>** | **62 (8.7)<sup>B</sup>** | 15 (2.1) | 89 (12.6) | 91 (12.8) | 14 (2.0) | 13 (1.8) |
| Female (B) | 577 | 315 (54.6) | 12 (2.0) | 9 (1.6) | 39 (6.8) | 14 (2.4) | 82 (14.2) | 65 (11.3) | 23 (4.0) | 18 (3.1) |
| Other (C) | 7 | **5 (71.4)<sup>A,B</sup>** | 0 | 0 | 0 | 0 | 1 (14.3) | 0 | 1 (14.3) | 0 |
| **Access to Dental Insurance** |  |  |  |  |  |  |  |  |  |  |
| Yes (D) | 373 | 200 (53.6) | 8 (2.1) | 9 (2.4) | 25 (6.7) | 7 (1.9) | 59 (15.8) | 44 (11.8) | 14 (3.8) | 7 (1.9) |
| No (E) | 923 | 493 (53.4) | 31 (3.4) | 28 (3.0) | 76 (8.3) | 23 (2.5) | 113 (12.2) | 112 (12.1) | 23 (2.5) | 24 (2.6) |
| **Age** |  |  |  |  |  |  |  |  |  |  |
| 18-34 (F) | 228 | 75 (32.9) | **13 (5.7)<sup>I</sup>** | 11 (4.8) | 17 (7.5) | **11 (4.8)<sup>H</sup>** | **47 (20.6)<sup>H,I</sup>** | **37 (16.2)<sup>I</sup>** | **14 (6.2)<sup>I</sup>** | 3 (1.3) |
| 35-44 (G) | 183 | 83 (45.4) | 5 (2.7) | 2 (1.1) | 16 (8.8) | **9 (4.9)<sup>H</sup>** | **31 (16.9)<sup>I</sup>** | 24 (13.1) | **6 (3.3)<sup>I</sup>** | 7 (3.8) |
| 45-64 (H) | 526 | **283 (53.8)<sup>F,G</sup>** | **16 (3.0)<sup>I</sup>** | 16 (3.0) | **49 (9.4)<sup>I</sup>** | 9 (1.7) | **69 (13.1)<sup>I</sup>** | 60 (11.4) | **15 (2.9)<sup>I</sup>** | 9 (1.7) |
| 65+ (I) | 358 | **252 (70.4)<sup>F,G,H</sup>** | 5 (1.4) | 8 (2.2) | 19 (5.3) | 0 | 24 (6.7) | 35 (9.8) | 2 (0.6) | 13 (3.6) |
| **Education** |  |  |  |  |  |  |  |  |  |  |
| High school or less (J) | 327 | 170 (52.0) | 9 (2.8) | 9 (2.8) | 29 (8.9) | 6 (1.8) | 38 (11.6) | 48 (14.7) | 11 (3.3) | 7 (2.1) |
| College/Trades (K) | 543 | 287 (52.9) | 20 (3.7) | 19 (3.5) | 42 (7.7) | 13 (2.4) | 69 (12.7) | 61 (11.3) | 16 (2.9) | 16 (2.9) |
| University (L) | 420 | 234 (55.7) | 10 (2.4) | 9 (2.1) | 30 (7.1) | 11 (2.6) | 64 (15.3) | 46 (11.0) | 8 (1.9) | 8 (1.9) |
| **Employment status** |  |  |  |  |  |  |  |  |  |  |
| Employed (M) | 628 | 274 (43.7) | **30 (4.8)<sup>O</sup>** | 21 (3.3) | **56 (8.9)<sup>O</sup>** | **22 (3.5)<sup>O</sup>** | **106 (16.9)<sup>O</sup>** | **90 (14.3)<sup>O,P</sup>** | 12 (1.9) | 17 (2.7) |
| Unemployed (looking for work) (N) | 72 | 39 (54.1) | 0 | 4 (5.6) | 4 (5.6) | **5 (6.9)<sup>O</sup>** | 8 (11.1) | 9 (12.5) | 3 (4.2) | 0 |
| Not employed/not in workforce (O) | 556 | **364 (65.5)<sup>M,N</sup>** | 8 (1.4) | 12 (2.2) | 35 (6.3) | 3 (0.5) | 51 (9.2) | 54 (9.7) | 15 (2.7) | 14 (2.5) |
| Other (P) | 29 | 13 (44.9) | 0 | 0 | 5 (17.2) | 0 | 5 (17.2) | 1 (3.5) | **5 (17.2)<sup>M,O</sup>** | 0 |
| **Marital status** |  |  |  |  |  |  |  |  |  |  |
| Never Married (Q) | 405 | 221 (54.6) | 7 (1.7) | 9 (2.2) | 29 (7.2) | 5 (1.2) | 55 (13.6) | 51 (12.6) | **19 (4.7)<sup>R</sup>** | 9 (2.2) |
| Married or Common-Law (R) | 601 | 310 (51.6) | **27 (4.5)<sup>Q,S</sup>** | 22 (3.7) | 48 (8.0) | **19 (3.2)<sup>Q</sup>** | 78 (13.0) | 74 (12.3) | 10 (1.6) | 13 (2.1) |
| Divorced/separated or Widowed (S) | 286 | 161 (56.3) | 5 (1.7) | 6 (2.1) | 24 (8.4) | 5 (1.7) | 38 (13.3) | 30 (10.5) | 8 (2.8) | 9 (3.2) |
| **Parent of children** |  |  |  |  |  |  |  |  |  |  |
| <12 years (T) | 187 | 59 (31.6) | 9 (4.8) | 7 (3.7) | 19 (10.2) | **10 (5.3)<sup>W</sup>** | **39 (20.9)<sup>V,W</sup>** | 33 (17.6) | **9 (4.8)<sup>V</sup>** | 2 (1.1) |
| Age 12-17 (U) | 200 | 77 (38.5) | 11 (5.5) | 4 (2.0) | 22 (11.0) | 8 (4.0) | **37 (18.5)<sup>W</sup>** | 29 (14.5) | 5 (2.5) | 7 (3.5) |
| 18 or older (V) | 226 | **121 (53.5)<sup>T,U</sup>** | 12 (5.3) | 11 (4.9) | 19 (8.4) | 6 (2.7) | 28 (12.4) | 24 (10.6) | 2 (0.9) | 3 (1.3) |
| No children (W) | 777 | **466 (60.0)<sup>T,U</sup>** | 17 (2.2) | 17 (2.2) | 53 (6.8) | 11 (1.4) | 82 (10.6) | 87 (11.2) | **24 (3.0)<sup>V</sup>** | 20 (2.6) |
| **Regions of Canada** |  |  |  |  |  |  |  |  |  |  |
| New Brunswick; Newfoundland and Labrador; Nova Scotia, Prince Edward Island (Atlantic Canada) (X) | 200 | 100 (50.0) | 7 (3.5) | 5 (2.5) | 20 (10.0) | 1 (0.5) | 30 (15.0) | 29 (14.5) | 5 (2.5) | 3 (1.5) |
| Quebec (Y) | 279 | 153 (54.8) | **5 (1.8)<sup>AC</sup>** | 9 (3.2) | 24 (8.6) | 6 (2.2) | 37 (13.3) | 35 (12.5) | 0 | 10 (3.6) |
| Ontario (Z) | 287 | 149 (51.9) | **12 (4.2)<sup>AC</sup>** | 10 (3.5) | 20 (7.0) | **11 (3.8)<sup>X</sup>** | 32 (11.2) | 36 (12.5) | 11 (3.8) | 6 (2.1) |
| Manitoba and Saskatchewan (AA) | 200 | 101 (50.5) | **8 (4.0)<sup>AC</sup>** | 6 (3.0) | 15 (7.5) | 4 (2.0) | **35 (17.5)<sup>Z</sup>** | 22 (11.0) | 6 (3.0) | 3 (1.5) |
| Alberta (AB) | 155 | 84 (54.2) | 3 (1.9) | 5 (3.2) | 8 (5.2) | 3 (1.9) | 27 (17.4) | 17 (11.0) | 6 (3.9) | 2 (1.3) |
| British Columbia, Yukon, Northern Territories and Nunavut (BC/North) (AC) | 201 | **116 (57.7)<sup>X</sup>** | 4 (2.0) | 1 (0.5) | 18 (9.0) | 0 | 26 (12.9) | 21 (10.4) | 9 (4.5) | 6 (3.0) |
| **Area of residence** |  |  |  |  |  |  |  |  |  |  |
| Urban (AD) | 1,109 | 591 (53.3) | 32 (2.9) | 32 (2.9) | 92 (8.3) | **24 (2.1)<sup>AE</sup>** | 156 (14.1) | 134 (12.1) | 31 (2.8) | 17 (1.5) |
| Rural (AE) | 171 | 87 (50.9) | 6 (3.5) | 3 (1.7) | 11 (6.4) | 1 (0.6) | 27 (15.8) | 22 (12.9) | 5 (2.9) | **9 (5.3)<sup>AD</sup>** |
| Remote (AF) | 41 | 25 (61.0) | 1 (2.4) | 1 (2.4) | 2 (4.8) | 0 | 4 (9.8) | 4 (9.8) | 0 | 4 (9.8) |

\*CRA: Canada Revenue Agency
Letters in uppercase indicate comparisons within the same column with a *p*-value <0.05

Associations between participants’ support for the CDCP and covariates are reported in Table 4. Overall, 81.5% indicated their support the CDCP (58.2% strongly support, 23.3% somewhat support) while 3.5% opposed the program. Those without access to dental insurance were significantly more likely to support the CDCP than those with insurance (83.6% vs. 76.4%). Additionally, those with university education were more likely to support the program than those with ≤ high school or those who completed college/trades. Respondents from Atlantic Canada and British Columbia, Yukon, Northwest Territories, and Nunavut (BC/North) regions were more likely to support the CDCP, while the lowest support came from the Manitoba and Saskatchewan region and Alberta. No statistical differences in support were observed between those living in urban, rural, or remote regions of Canada.

**Table 4.** Support or Oppose CDCP.

|  | **Total** | **Support (total)** | Strongly support | Somewhat support | **Oppose (total)** | Strongly oppose | Somewhat oppose | **Neither support nor oppose** | **Don’t know** |
|---|---|---|---|---|---|---|---|---|---|
| **Total overall** | 2,220 | 1,808 (81.5) | 1,291 (71.4) | 517 (28.6) | 79 (3.6) | 39 (49.4) | 40 (50.6) | 234 (10.5) | 97 (4.4) |
| **Gender** |  |  |  |  |  |  |  |  |  |
| Male (A) | 1,056 | 860 (81.4) | 602 (57.0) | **258 (24.4)<sup>C</sup>** | 46 (4.4) | 21 (2.0) | **25 (2.4)<sup>B</sup>** | 115 (10.9) | 35 (3.3) |
| Female (B) | 1,143 | 935 (81.8) | 679 (59.4) | **256 (22.4)<sup>C</sup>** | 32 (2.8) | 18 (1.6) | 14 (1.2) | 116 (10.2) | **60 (5.3)<sup>A</sup>** |
| Other (C) | 16 | 11 (68.8) | 10 (62.5) | 1 (6.3) | 1 (6.3) | 0 | 1 (6.3) | 2 (12.5) | 2 (12.5) |
| **Access to Dental Insurance** |  |  |  |  |  |  |  |  |  |
| Yes (D) | 674 | 515 (76.4) | 341 (50.6) | 174 (25.8) | 31 (4.6) | 17 (2.5) | 14 (2.1) | **94 (14.0)<sup>E</sup>** | 34 (5.0) |
| No (E) | 1,547 | **1,294 (83.6)<sup>D</sup>** | **951 (61.5)<sup>D</sup>** | 343 (22.2) | 49 (3.2) | 22 (1.4) | 27 (1.7) | 141 (9.1) | **63 (4.1)<sup>D</sup>** |
| **Age** |  |  |  |  |  |  |  |  |  |
| 18-34 (F) | 494 | 416 (84.2) | 254 (51.4) | **162 (32.8)<sup>G,H,I</sup>** | 6 (1.2) | 2 (0.4) | 4 (0.8) | 49 (9.9) | 23 (4.7) |
| 35-44 (G) | 335 | 270 (80.6) | 186 (55.5) | 84 (25.1) | **17 (5.1)<sup>F</sup>** | 5 (1.5) | **12 (3.6)<sup>H,I</sup>** | 34 (10.2) | 14 (4.2) |
| 45-64 (H) | 832 | 671 (80.7) | **508 (61.1)<sup>F</sup>** | 163 (19.6) | **38 (4.6)<sup>F</sup>** | **20 (2.4)<sup>F</sup>** | 18 (2.2) | 85 (10.2) | 38 (4.6) |
| 65+ (I) | 557 | 451 (81.0) | **343 (61.6)<sup>F</sup>** | 108 (19.4) | 17 (3.1) | 11 (2.0) | 6 (1.1) | 67 (12.0) | 22 (4.0) |
| **Education** |  |  |  |  |  |  |  |  |  |
| High School or less (J) | 728 | 586 (80.5) | 434 (59.6) | 152 (20.9) | 17 (2.3) | 9 (1.2) | 8 (1.1) | 77 (10.6) | **48 (6.6)<sup>K,L</sup>** |
| College/Trades (K) | 858 | 685 (79.8) | 470 (54.8) | 215 (25.1) | **38 (4.4)<sup>J</sup>** | 18 (2.1) | 20 (2.3) | 101 (11.8) | 34 (4.0) |
| University (L) | 626 | **532 (85.0)<sup>J,K</sup>** | **381 (60.9)<sup>K</sup>** | 151 (24.1) | 24 (3.8) | 12 (1.9) | 12 (1.9) | 55 (8.8) | 15 (2.4) |
| **Employment Status** |  |  |  |  |  |  |  |  |  |
| Employed (M) | 1,066 | 869 (81.5) | 587 (55.1) | **282 (26.5)<sup>O,P</sup>** | **45 (4.2)<sup>N</sup>** | 21 (2.0) | **24 (2.3)<sup>N</sup>** | 115 (10.8) | 37 (3.5) |
| Unemployed/looking for work (N) | 178 | 150 (84.3) | 112 (62.9) | 38 (21.3) | 3 (1.7) | 2 (1.1) | 1 (0.6) | 14 (7.9) | 11 (6.2) |
| Not employed/not in workforce (O) | 908 | 739 (81.4) | **553 (60.9)<sup>M</sup>** | 186 (20.5) | 26 (2.9) | 15 (1.7) | 11 (1.2) | 98 (10.8) | 45 (5.0) |
| Other (P) | 52 | 40 (76.9) | 34 (65.4) | 6 (11.5) | 5 (9.6) | 0 | **5 (9.6)<sup>N,O</sup>** | 5 (9.6) | 2 (3.8) |
| **Marital Status** |  |  |  |  |  |  |  |  |  |
| Never Married (Q) | 807 | 657 (81.4) | 449 (55.6) | **208 (25.8)<sup>S</sup>** | 26 (3.2) | 11 (1.4) | 15 (1.9) | 85 (10.5) | 39 (4.8) |
| Married/Common-law (R) | 977 | 791 (81.0) | 551 (56.4) | **240 (24.6)<sup>S</sup>** | 35 (3.6) | 17 (1.7) | 18 (1.8) | 108 (11.1) | 43 (4.4) |
| Divorced/separated or widowed (S) | 430 | 357 (83.0) | **288 (67.0)<sup>Q,R</sup>** | 69 (16.0) | 17 (4.0) | 10 (2.3) | 7 (1.6) | 42 (9.8) | 14 (3.3) |
| **Parent of children** |  |  |  |  |  |  |  |  |  |
| Age < 12 years (T) | 247 | 203 (82.2) | 133 (53.8) | 70 (28.3) | 7 (2.8) | 2 (0.8) | 5 (2.0) | 25 (10.1) | **12 (4.9)<sup>U</sup>** |
| Age 12-17 (U) | 253 | 214 (84.6) | 156 (61.7) | 58 (22.9) | **15 (5.9)<sup>T,V</sup>** | 6 (2.4) | **9 (3.6)<sup>V</sup>** | 20 (7.9) | 4 (1.6) |
| Age ≥18 (V) | 382 | 312 (81.7) | 228 (59.7) | 84 (22.0) | 11 (2.9) | 7 (1.8) | 4 (1.0) | **52 (13.6)<sup>U</sup>** | 7 (1.8) |
| No children (W) | 1,435 | 1,164 (81.1) | 834 (58.1) | 330 (23.0) | 49 (3.4) | 25 (1.7) | 24 (1.7) | 149 (10.4) | **73 (5.1)<sup>V,W</sup>** |
| **Regions of Canada** |  |  |  |  |  |  |  |  |  |
| New Brunswick; Newfoundland and Labrador; Nova Scotia, Prince Edward Island (Atlantic Canada) (X) | 323 | **287 (88.9)<sup>Y,Z,AA,AB</sup>** | **226 (7.0)<sup>Y,Z,AA,AB</sup>** | 61 (18.9) | 10 (3.1) | 7 (2.2) | 3 (0.9) | 18 (5.6) | 8 (2.5) |
| Quebec (Y) | 475 | **399 (84.0)<sup>AA,AB</sup>** | **281 (59.2)<sup>AB</sup>** | **118 (24.8)<sup>X</sup>** | 10 (2.1) | 5 (1.1) | 5 (1.1) | **47 (9.9)<sup>X</sup>** | 19 (4.0) |
| Ontario (Z) | 498 | 397 (79.7) | 279 (56.0) | 118 (23.7) | 17 (3.4) | 5 (1.0) | 12 (2.4) | **60 (12.0)<sup>X,AC</sup>** | 24 (4.8) |
| Manitoba and Saskatchewan (AA) | 284 | 214 (75.4) | 149 (52.5) | 65 (22.9) | **21 (7.4)<sup>Y,Z,AC</sup>** | **14 (4.9)<sup>Y,Z,AC</sup>** | 7 (2.5) | **37 (13.0)<sup>X,AC</sup>** | 12 (4.2) |
| Alberta (AB) | 306 | 237 (77.5) | 158 (51.6) | **79 (25.8)<sup>X</sup>** | **17 (5.6)<sup>Y,AC</sup>** | **11 (3.6)<sup>Y,Z,AC</sup>** | 6 (2.0) | **38 (12.4)<sup>X</sup>** | 14 (4.6) |
| British Columbia, Yukon, Northern Territories and Nunavut (BC/North) (AC) | 334 | **285 (85.3)<sup>Z,AA,AB</sup>** | **216 (64.7)<sup>Z,AA,AB</sup>** | 69 (20.7) | 8 (2.4) | 4 (1.2) | 4 (1.2) | 26 (7.8) | 15 (4.5) |
| **Area of residence** |  |  |  |  |  |  |  |  |  |
| Urban (AD) | 1,829 | 1,497 (81.8) | 1,059 (57.9) | 438 (23.9) | 66 (3.6) | 37 (2.0) | 29 (1.6) | 190 (10.4) | 76 (4.2) |
| Rural/Remote (AE) | 390 | 321 (82.3) | 249 (63.8) | 72 (18.5) | 17 (4.6) | 9 (2.3) | 8 (2.1) | 36 (9.2) | 16 (4.1) |

Total is comprised of **support (total)**, **oppose (total)**, **neither support or oppose** and **don’t know** categories. Sub-categories (e.g. **strongly support**, **strongly oppose**) percentages were calculated separately
Letters in uppercase indicate comparisons within the same column with a *p*-value <0.05

Primary reasons (hereby labelled as “motivators”) for why participants would apply for the CDCP are summarized on Figure 1. The most common motivators included the extent of coverage offered by the CDCP, the prospect of better personal dental health, if the future enrolment process would be simple, and if they would have a good understanding of the plan. The distribution of motivators to apply for the CDCP by demographic characteristics are presented in Figures 2A and 2B. Among all the evaluated sociodemographic variables, participants without dental insurance were the most motivated/with the most significant number of motivators to apply for the CDCP. Respondents with an education level lower than high school were significantly more motivated to apply than all others if they lived closer to a dentist or oral health professional (47.4%). Additionally, they were also statistically more motivated to apply if the program was recommended by their healthcare provider (38.9%), when compared to participants with a university level of education (24.5%). Parents aged 35-44 were the least interested to apply for most motivators, except children’s urgent dental needs (38.4%), more than one eligible child requiring dental care (43.0%), and whether the program was recommended by their child’s healthcare provider (30.6%).

**Figure 1.**
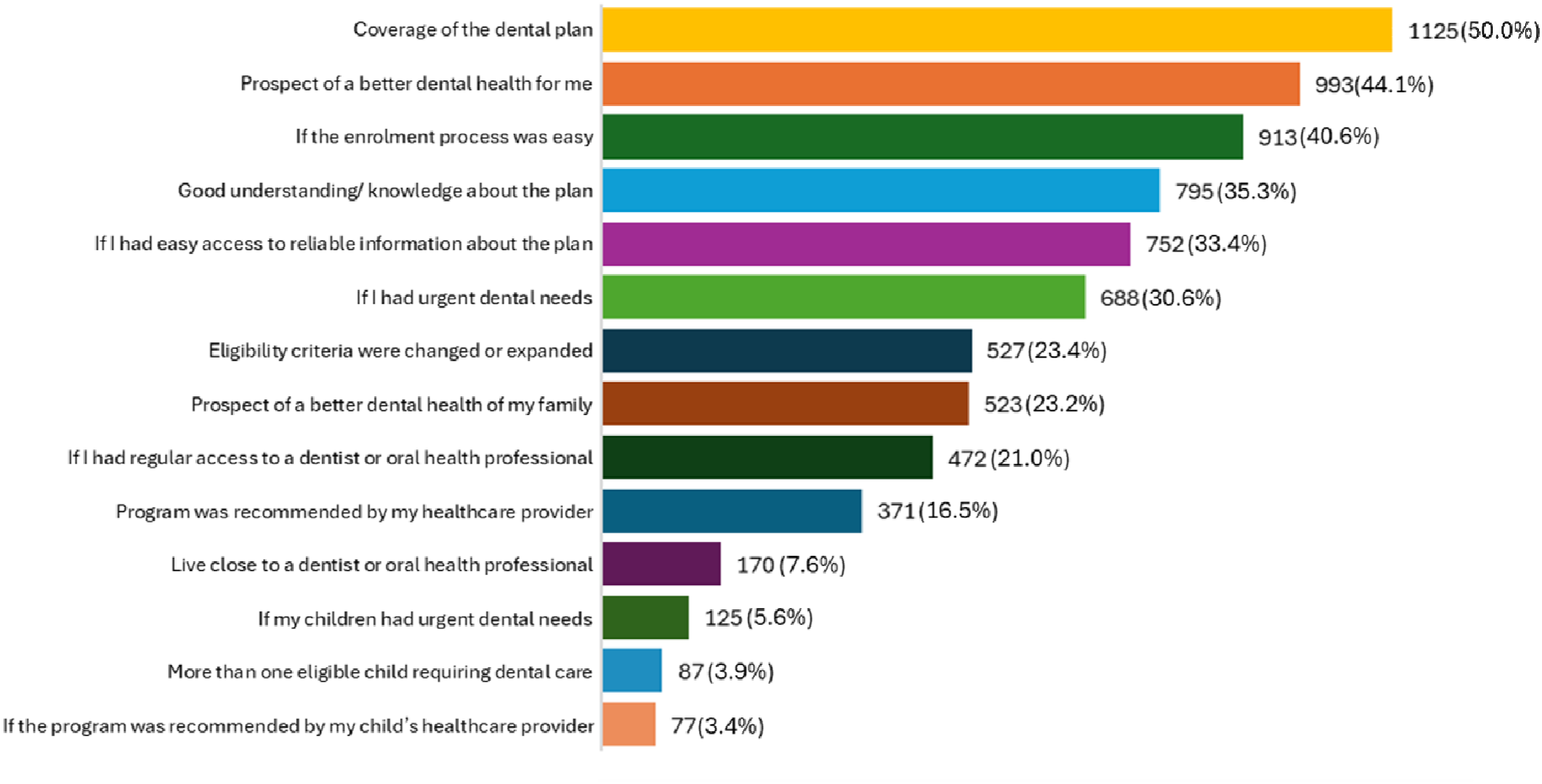
Motivators to apply for the CDCP

**Figure 2.**
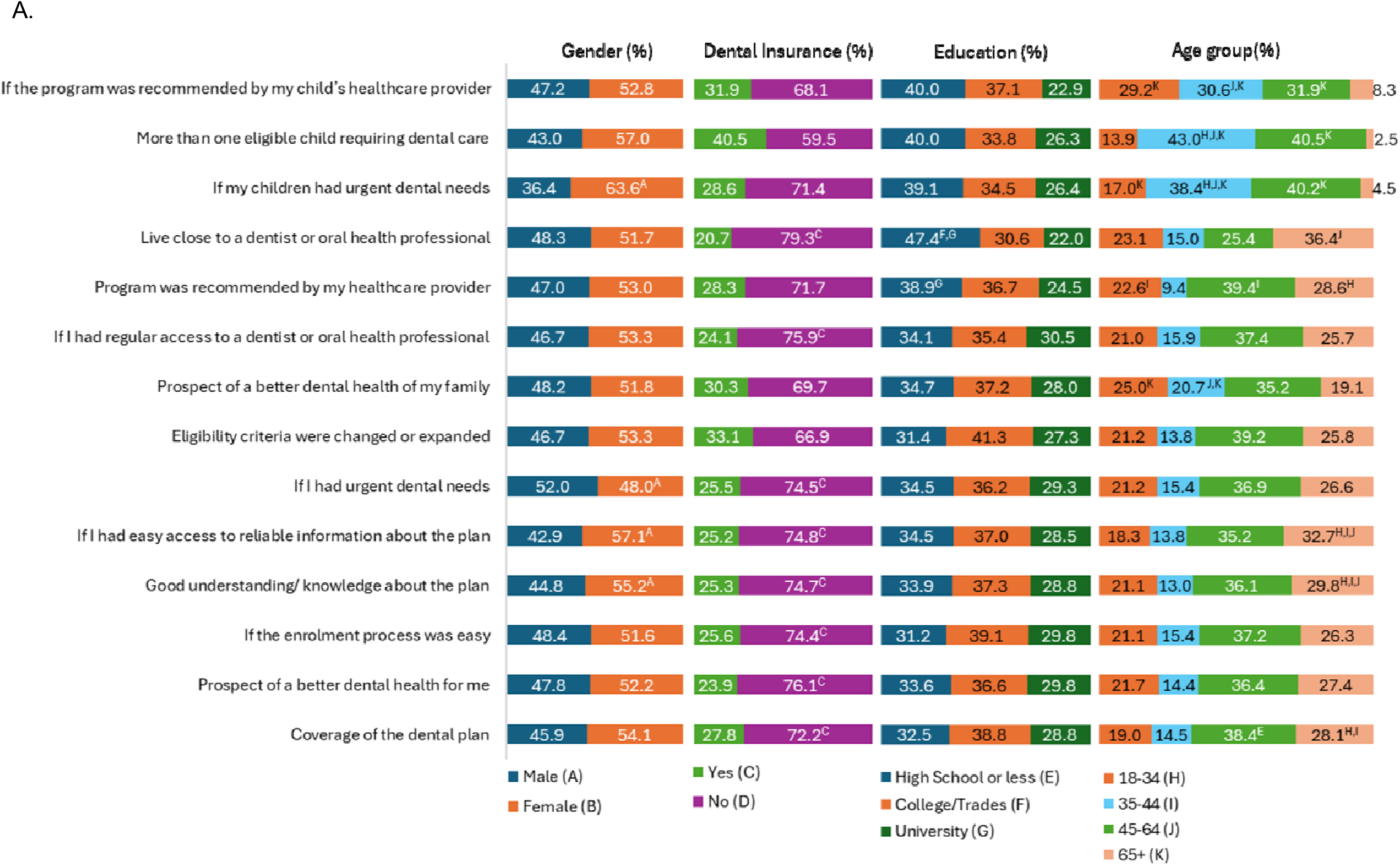
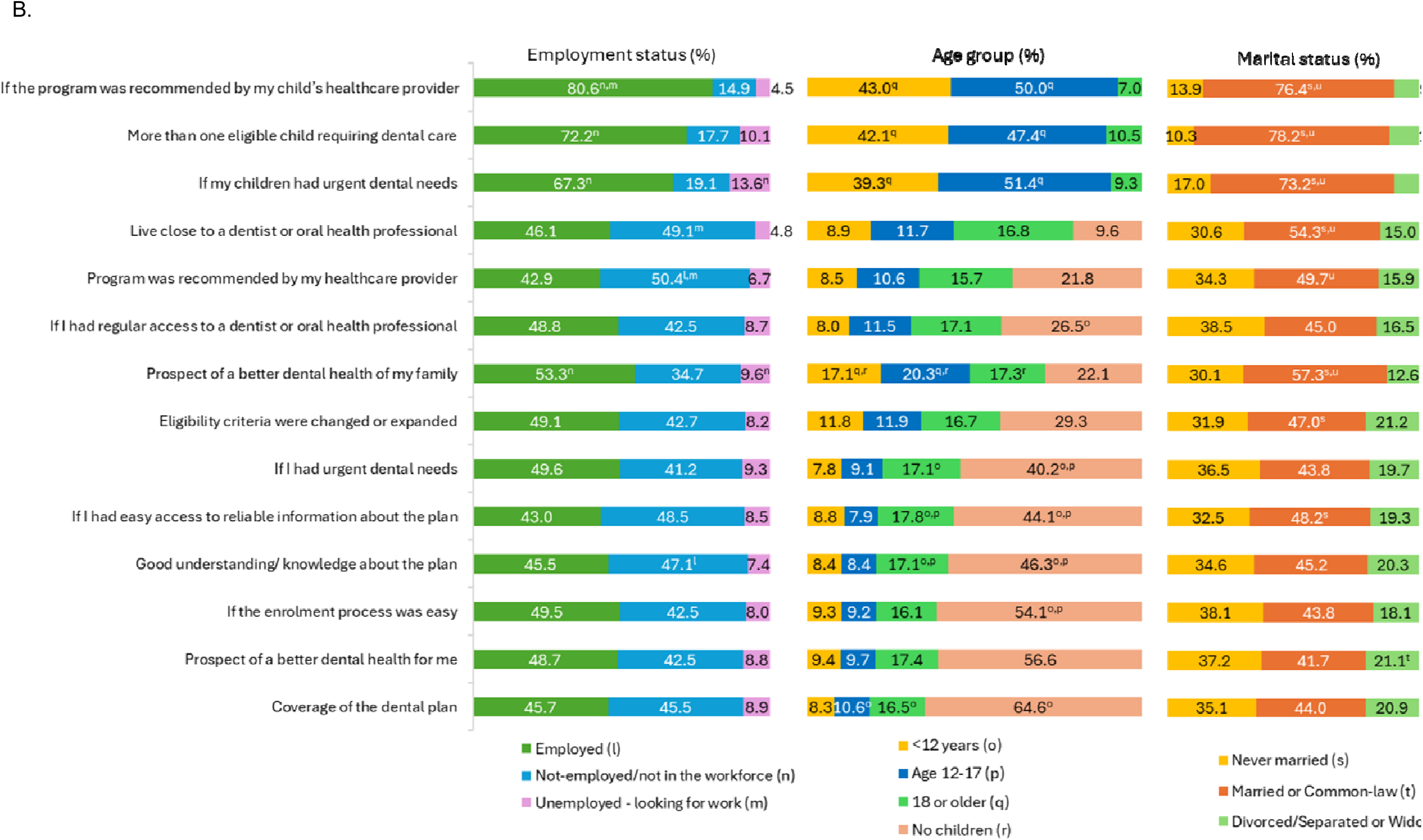
Motivators to apply for the CDCP by sociodemographic variables Letters in uppercase indicate comparisons with a *p*-value <0.05

Married (or in common-law) participants were significantly more motivated to apply for the CDCP, when compared to those never married and those divorced or separated. Married/common-law participants were significantly more motivated if they had more than one child in need of dental care (78.2%), if the program was recommended by their child’s healthcare provider (76.4%), if their child had urgent dental needs (73.2%), and by the prospect of better dental health for their family (57.3%). Respondents who were employed were significantly more motivated to apply depending on their children’s urgent needs (67.3%), if they had more than one eligible child requiring dental care (72.2%) and whether the CDCP was recommended by their child’s healthcare provider (80.6%). Lastly, Figure 3 portrays the comparisons for motivators considering which regions of Canada in which respondents resided. Those from BC/North were statistically more inclined to apply for the CDCP than other regions for several motivators, such as the coverage of the dental plan itself (58.1%), whether the enrollment process was easy (50.0%) and if they had a good understanding of the plan (44.3%).

**Figure 3.**
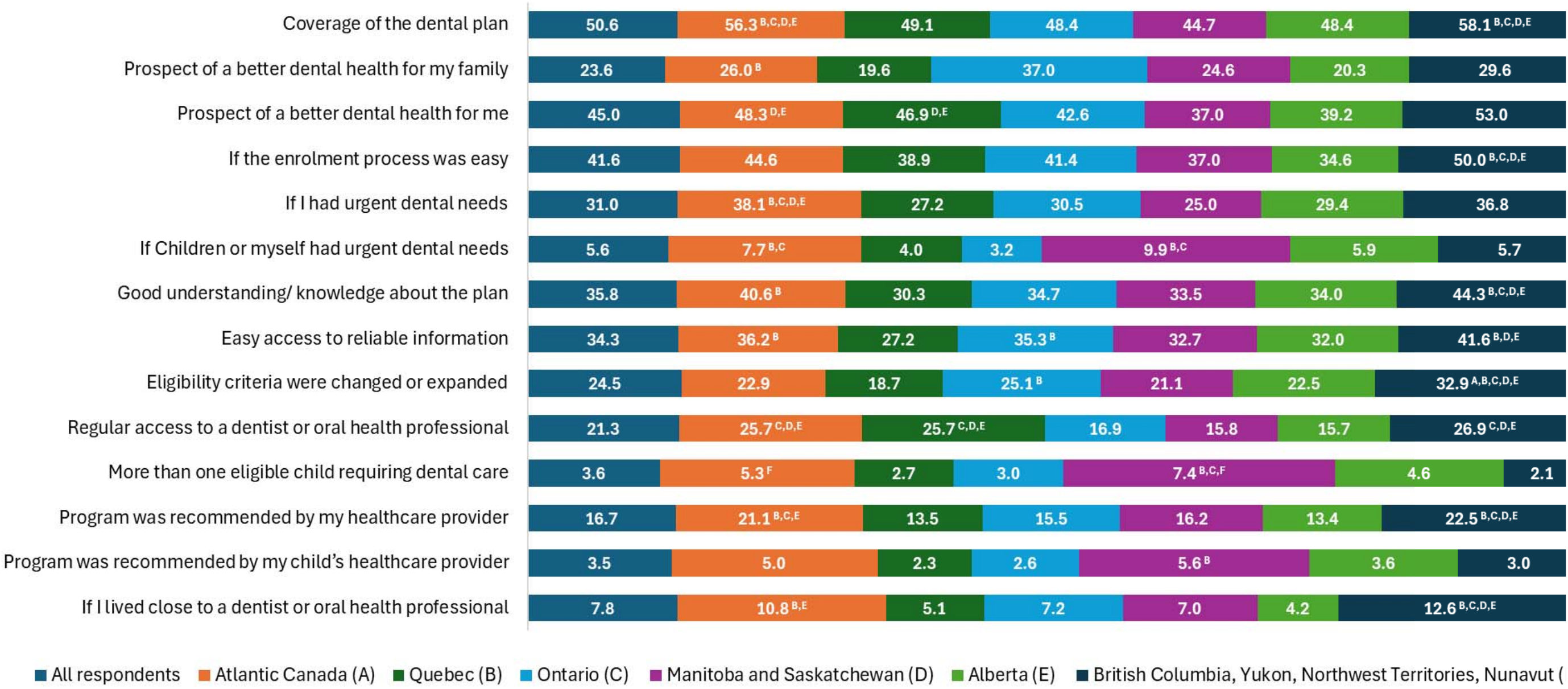
Motivators to apply for the CDCP by regions Letters in uppercase indicate comparisons with a *p*-value <0.05

## 4. Discussion

This study identified that, although only a moderate proportion of Canadian adults had heard of the CDCP before its official launch in 2024, their support for the new program was strong. The level of awareness of the CDCP at the time of the study can easily be explained as this survey was conducted while the government was planning the program and very little information about the CDCP was reaching the public or oral health professionals. Awareness of the CDCP was significantly higher among males, seniors (≥ 65 years of age), unemployed, married or in common-law relationships and those with higher education (College or University). Additionally, respondents from Atlantic Canada and Manitoba and Saskatchewan were significantly more aware than those from Alberta. Surprisingly, the absence of dental insurance and differences between urban and rural residence were not associated with having heard of the upcoming CDCP. Since the CDCP launch in 2024 awareness has undoubtedly increased in the general population^12^, but low awareness likely still remains among some priority populations that struggle to access oral health care, including First Nations, Métis, and Inuit peoples, those living in rural and remote parts of Canada, those already receiving provincial or federal government dental benefits, and newcomers to Canada.

Participants in this study indicated that they obtained their information about the CDCP from the news/media. Surprisingly, very few had learned of the CDCP from oral health practitioners. This may be because at the time this survey took place, details of the upcoming CDCP were not yet shared with oral health professionals. Current data from the Government of Canada reveal that over 29,200 oral health professionals nationwide are now participating in the CDCP^13^, and their position as source of information about the plan may be much more impactful nowadays.

An interesting finding was that respondents with children <12 years of age were more likely to search for information about the CDCP. One explanation for this may be because the Interim CDB, which was the precursor to the CDCP from October 2022 through June 2024 was limited to children <12 years of age.^8, 14–17^ Parents of children < 12 years of age may have already applied for the Interim CDB and may have had a heightened interest in obtaining more information about the upcoming CDCP.

Awareness of the CDCP also differed across age and education groups. Seniors ≥ 65 years of age were more likely to have heard about the CDCP than younger respondents. This is likely due to the fact that the first eligible group for the CDCP were senior citizens, and early government communications about the program targeted older adults. These findings emphasize the urgent need for targeted public education and advertising strategies, as some specific groups might need and benefit from the program the most. Menon *et al.,* in a study evaluating the CDCP outreach to senior populations, highlights that public awareness is an important challenge to the effective implementation of the CDCP.^18^ Health literacy is critical to the ability to use information and access the oral health services appropriately, since adults are the primary caregivers for both children and older adults, likewise increasing community participation.^19, 20^

Despite the relatively low reported awareness, the majority of participants (81.5%) supported the government intervention. Public support varied across Canada, with the lowest support coming from the prairie provinces, which is surprising since residents of these two provinces reported, the greatest access to oral health care barriers.^11^ That same study with a national survey of parents of children <12 years of age reported that 87% supported the Interim CDB.^11^ At the launch of the CDCP, a survey of Manitoba Dental Association members revealed opposition towards the program with 89% of dentists indicating that they were unlikely or very unlikely to participate in the CDCP.^11, 21^ Over the past two years, dentists’ acceptance of the CDCP has dramatically shifted, as over 90% of dentists in Canada are currently participating in the program.^22^ According to the Government of Canada official report, over 4.5 million applicants were approved to receive care since the deployment of the CDCP, as of June 30, 2026.^13^ Future studies will monitor both public and oral health professionals’ support for the CDCP, as a measure to assess whether the program is meeting the expectations of Canadian families.

Motivational factors, among others, play a role in influencing individual’s ability to make decisions.^23^ The main motivator for applying for the CDCP was dental coverage itself. However, even with the CDCP facilitating access to oral health care in Canada, the cost of dental care services remains a significant barrier for those individuals who are left with the remaining 40% or 60% co-pay, as those with adjusted AFNI < $70,000 are eligible for 100% coverage. Further, many private dental offices may balance-bill the difference between the CDCP and different provincial dental association fees. Some studies show that the Interim CDB was insufficient to fully cover all the dental procedures costs.^8, 15^ Overall, 40.6% of respondents indicated that a motivator to apply for the CDCP would be if the enrollment process was easy.

In addition, respondents emphasized a clear understanding of the plan, and access to reliable information as motivators to apply, reinforcing the need for accessible, and user-friendly communication strategies. A recent report from the Government of Canada with information from the first year of the CDCP showed that only 55% of the estimated eligible children of under 18 years of age were actually enrolled.^12^ Policy makers should invest in more effective, targeted communication for the general population and underserved groups, as the latter are those who would benefit the most from enrolling into the program. These crucial aspects must be incorporated into the CDCP education to promote more equitable access to healthcare and foster a culture of well-informed citizens who are able to make better decisions regarding their own health.^20^

This study is not without limitations. This survey was conducted from July to August 2023, while the CDCP was still under development, so the views of respondents likely have changed since then. Fortunately, these results serve as an important baseline of the public’s support and views on the program. Future studies should examine whether awareness of the CDCP increased after the program was launched, since the survey was conducted before implementation. Repeating the survey would help determine whether public communication and outreach improved awareness and whether some groups continue to face barriers to learning about the program. Strengths of this study include the nationally representative nature of the sample and the robust sample size. However, the limited number of parents of children < 18 years of age is a limitation.

## 6. Conclusion

Despite the low awareness reported by the participants, support for the CDCP was strong. These findings highlight the need for targeted advertising strategies to increase awareness and ensure that information reaches those most in need of the program. These findings also showcase how Canadian families seemed to be eager for the possibility of a publicly funded dental coverage. Follow-up studies are essential to evaluate the program’s reach, development and uptake after its implementation.

## Data Availability

All data produced by the Government of Canada Publications are available online at: https://epe.bac-lac.gc.ca/100/200/301/pwgsc-tpsgc/por-ef/health/2024/014-23-e/014-23_Report.pdf

## Acknowledgements

This study was supported by Dr. Schroth’s Canadian Institutes of Health Research Applied Public Health Chair in Oral Health program of research – “Public Health Approaches to Improve Access to Oral Health Care and Oral Health Status for Young Children in Canada”.

